# Validation of the new pathology staging system for progressive supranuclear palsy

**DOI:** 10.1101/2021.01.18.21250017

**Authors:** Mayen Briggs, Kieren SJ Allinson, Maura Malpetti, Maria Grazia Spillantini, James Benedict Rowe, Sanne Simone Kaalund

## Abstract

Progressive supranuclear palsy (PSP) is a neurodegenerative disorder associated with neuroglial accumulation of 4-repeat tau protein. Kovacs et al. have recently proposed a new semi-quantitative staging system to categorise the severity of PSP pathology, using the distribution of tau aggregates as it progresses from subcortical to cerebellar and cortical regions. Here, we test the new PSP pathology staging system in an independent series of PSP, and test the potential association between pathology stage and clinical severity at death. We include tissue from 35 people with a clinical diagnosis of PSP (including N=25 with Richardson’s syndrome and N=10 with other phenotypes). Donors had attended longitudinal clinical studies at the Cambridge Centre Parkinson-plus including assessment of clinical severity by the PSP rating scale (PSPRS) and cognitive performance by the revised Addenbrooke’s Cognitive Examination (ACE-R). We rated tau pathology from none-to-severe in six regions. We focused on (I) astrocytic tau inclusions in striatum, frontal and occipital regions, and (II) neuronal and oligodendroglia tau inclusions in globus pallidus, subthalamic nucleus, and cerebellum. Thirty-two cases (91%) readily conformed to the new staging system, ranging from stage 2 to 6. Staging system applied to brains from people with different clinical phenotypes of PSP. Neuropathology stages correlated with clinical severity at death using both PSPRS and ACE-R, weighted for the interval between last assessment and donation. Our study supports the proposed sequential distribution of tau aggregates in PSP pathology, and the hypothesised relationship between clinical and neuropathological severity. For future studies, in order to standardise rating between centres, we propose a set of operational criteria for region-specific thresholds or tau burden, and a visual guide.

## Introduction

Progressive supranuclear palsy (PSP) is a severe neurodegenerative disorder resulting in diverse clinical phenotypes with restricted eye movements, akinetic-rigidity, falls, cognitive and behavioural deficits [11]. The neuropathological diagnosis of PSP is based on the presence of intracellular aggregates of 4-repeat (4R) tau in neurons and glia, in basal ganglia, cerebral cortex, cerebellar dentate nucleus and white matter [3, 10, 17, 27]. In addition to tufted astrocytes, the characteristic glial cytopathologies of PSP include thorn-shaped astrocytes, coiled bodies in oligodendrocytes, and white matter threads. The neuronal pathology comprises globose neuronal tangles, pre-tangles, and threads in subcortical nuclei and cortical grey matter. The pathology is hypothesised to progress from neurons and oligodendroglia in substantia nigra, globus pallidus and subthalamic nucleus to astrocytes of the precentral gyrus in the cerebral cortex and striatum, before reaching the neurons and oligodendrocytes of cerebellum and/or frontal cortex [30]. Later, the neuroglial pathology extends to frontal and occipital cortex [14].

The low prevalence of clinical PSP [2] and rarity of pre-symptomatic cases with PSP pathology [5, 6, 15, 22, 26, 31] makes the development and evaluation of a PSP pathology staging scheme challenging. Further, although the PSP-specific tau aggregates are primarily made of hyperphosphorylated 4R tau, neurofibrillary tangles and threads in PSP are virtually indistinguishable from the 3R/4R aggregates characterising Alzheimer’s disease when stained by conventional diagnostic antibodies against hyperphosphorylated tau (i.e. Gibbons et al. [8]). Thus, the incidence of concomitant tau pathology in PSP, Alzheimer’s disease and Argyrophilic grain disease (AGD) was not fully appreciated until more recent studies using antibodies specific for Alzheimer’s disease tangles (GT-38) [8, 12]. These studies revealed a high percentage of concomitant pathology across the spectrum of frontotemporal lobar degeneration [8], particularly in PSP where concomitant Alzheimer’s disease of Braak stage I-III [12, 20, 28] and primary age-related tauopathy (PART) [15] are frequent. The concomitant pathologies may not determine the clinical PSP syndrome or symptom severity [24], but they complicate the evaluation of tau pathology particularly in midbrain and pontine nuclei, where early PSP tau pathology is predominantly neuronal [14, 17, 30].

A new staging system was proposed in 2020 by Kovacs et al, describing a sequential distribution of PSP cytopathology [14]. Each stage is defined by the combination of semi-quantitative scores of specific cellular pathologies in six select regions. The system proposes a consistent and progressive pattern, from stage 1 towards stage 6. Importantly, the Kovacs *et al* system reduces reliance on regions with high risk of concomitant pathology and focusses on cell type specific tau-pathology.

This study tests two hypotheses. First, that the neuropathology stage correlates with clinical disease. This hypothesis builds on previous evidence that frequency and severity of *ante mortem* cognitive impairments is linked to *post mortem* total tau load [13, 24]. Second, that the staging system is applicable not only to Richardson’s syndrome cases but also to people with other PSP clinical phenotypes. The differential distributions of tau in clinical subtypes supports such a relationship [1, 4, 14, 16, 23, 25, 30]. To satisfy both these hypotheses would confirm the new system as an effective and reliable staging method [29].

We applied the new PSP pathology staging to a series of 35 new PSP cases including 25 cases with a clinical diagnosis of PSP-Richardson’s syndrome (PSP-RS) and 10 with other clinical variants of PSP. We investigated the correlations of pathology stage and regional cytology scores, with a*nte mortem* measures of disease severity and cognitive function, namely the PSP rating scale (PSPRS) and the revised Addenbrooke’s Cognitive Examination.

## Material and methods

### Case cohort

We include brain tissue donated to the Cambridge Brain Bank by 35 people with PSP who had participated in clinical studies at the Cambridge University Centre for Parkinson plus. Tissue was obtained under the ethically approved protocol for “Neurodegeneration Research in Dementia” (REC 16/WA/0240). Cases were selected based on a pathological diagnosis of PSP following the National Institute of Neurological Disorders and Stroke (NINDS) criteria, and availability of ante-mortem data including PSP rating scale [9] and the revised Addenbrooke’s Cognitive Examination (ACE-R)[21]. The number of clinical review visits per patient where PSPRS and/or ACE-R were recorded ranged from 1 to 13, with median of 4.

The clinical diagnoses included probable PSP-RS (n=25), probable PSP-frontal (n=1), possible PSP-corticobasal syndrome (n=3), possible PSP-progressive gait freezing (n=1), suggestive of PSP-corticobasal syndrome (n=3), suggestive of PSP-speech/language disorder (n=1) and suggestive of PSP-Parkinsonism (n=1). For patients dying before the publication of the revised criteria for PSP [11] the new criteria were applied to clinical data at the last clinic visit (details in Gazzina et al. 2019 [7]) (Table 1).

### Immunohistochemistry and pathological evaluation

In all cases neuropathological evaluation and staging was performed on the left hemisphere.

Following the proposed guidelines for PSP pathology staging [14] paraffin-embedded tissue blocks including the globus pallidus (GP), subthalamic nucleus (STN), striatum (STR), middle frontal gyrus (Fr), dentate nucleus and cerebellar white matter (DE/CB), and occipital cortex (Oc) were immunohistochemically stained for hyperhosphorylated tau, AT8 (AT8, MN1020, Thermo Scientific, USA). Cytopathologies were scored for the six regions on a 4 point scale from none (-) to severe (+++); *none* (-) was defined as no or single cell pathology for every 20^th^ field of view under a 40X objective. Specifically, we evaluated neuronal and oligodendroglial cytopathology in the GP and DE/CB, neuronal cytopathology in the STN, and astrocytic cytopathology in STR, Fr and Oc. If neurofibrillary tangles were observed in the STR in addition to astrocytic tau, a rating of moderate or severe was given. Neuronal cytopathology included neurofibrillary tangles, diffuse cytoplasmic inclusions and threads. Oligodendrocytic cytopathology included coiled bodies, while astrocytic cytopathology included tufted astrocytes and thorn-shaped astrocytes. Figure 1 provides a visual guide for the scoring system for each of the six regions.

**Fig. 1.**
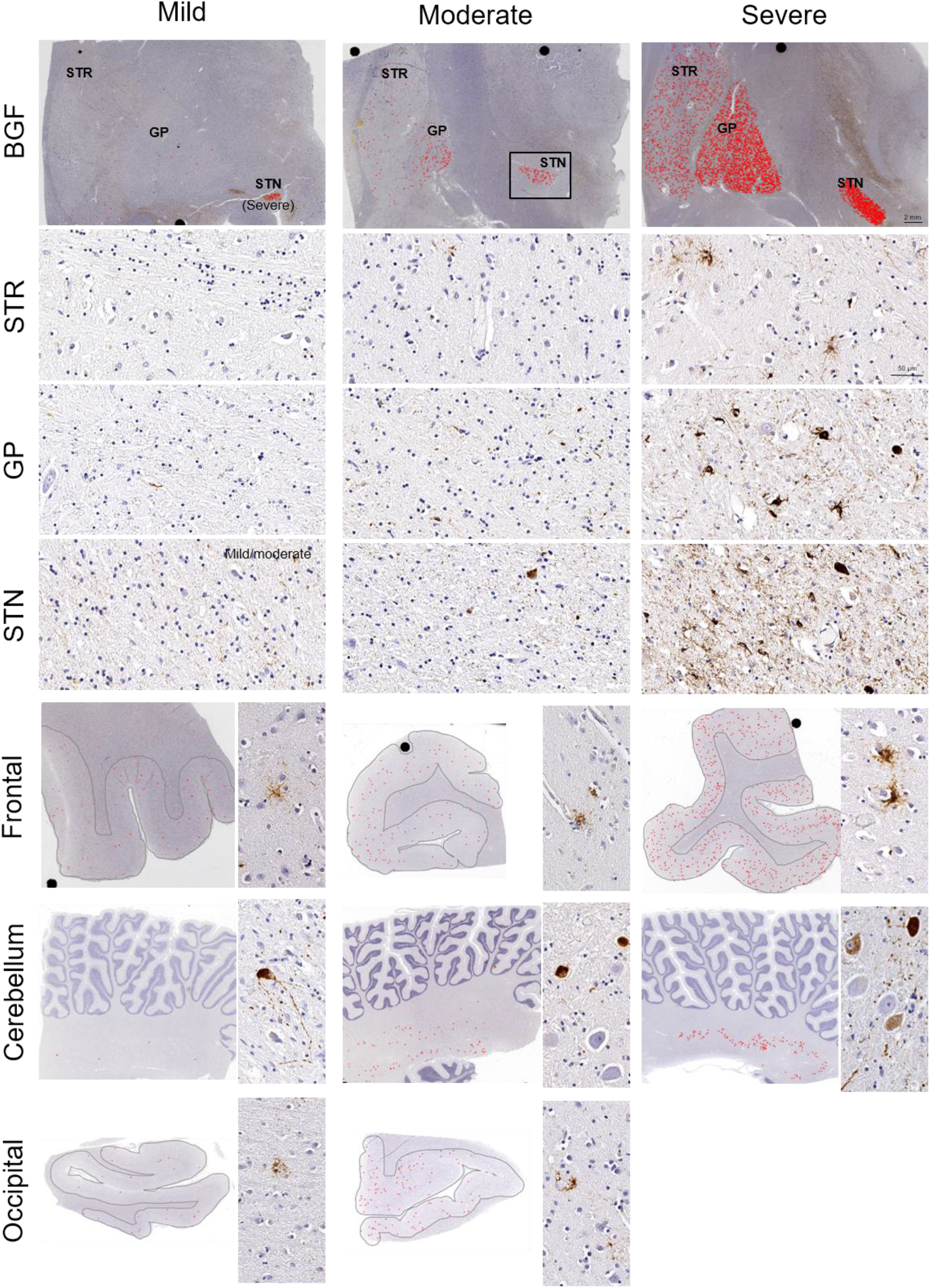
Visual guide to rating of PSP tau pathology. Micrographs of sections stained for hyperphosphorylated tau (AT8) showing the regional density of tau-aggregates (red points) at low magnification (0.3X, scale bar 2mm), and at higher magnification (20x, scale bar 50 μm). Left column shows regions rated “mild” – with the exception of STN where we did not have an example of “mild” pathology, thus the STN on the low magnification has “severe” pathology, and at high magnification we chose an area of STN with close to mild pathology from a STN rated “moderate”. The middle panel shows regions rated “medium” – on the low magnification, an example of “moderate” STN is superimposed on the STN for the given section which had a rating of “severe”. Right column shows regions rated “severe” – with the exception of occipital lobe as we did not have an example of “severe” occipital pathology, and the “severe” rating does not feature in the staging system. For cerebellum only pathology in the dentate nucleus, not white matter, is represented. Large black dots on the low magnification micrographs are image artefacts from the digital slide scanning. BGF – basal ganglia, STR – striatum, GP – globus pallidus, STN – subthalamic nucleus, Frontal – middle frontal gyrus, Occipital – primary visual cortex.

Tissue blocks for pathological diagnosis were sampled according to NINDS standard guidance for neurodegenerative diseases from brainstem, subcortical and cortical areas and were evaluated for the initial pathological diagnosis of PSP and possible concomitant pathologies of amyloid beta (Clone 6F/3D, M0872, Dako, Denmark), alpha-synuclein (SA3400, Enzo life sciences, USA) and TDP-43 (TIP-PTD-P02, Cosmo Bio Co LTD, Japan), and vascular pathology.

### Statistical analysis

Statistical differences in age at death and symptom duration were analysed by Kruskal Wallis test. Disease duration was calculated as time interval between symptom onset and death. Main effects of tau pathology scores on *ante mortem* clinical severity, as measured by the PSPRS and ACE-R scores at the last individual clinical visit, were analysed using one-way weighted means ANOVA. The reciprocal of time between testing and death was used as the weight, giving more weight to those data points with the shortest interval from test date to date of death. Then, we also imputed the PSPRS and ACE-R scores at death using multivariate imputation by chain equations as described in Malpetti et al. [19] (Supplementary figure 1 and 2). In brief, we used the individual longitudinal data for PSPRS and ACE-R scores, and months from baseline to each follow-up visit to estimate missing scores for disease severity (PSPRS) and cognition (ACE-R), including scores at death. Patient reference numbers were included in the imputation to account for individual differences. Four patients had no PSPRS assessment, and PSPRS scores was imputed from the group mean adjusted for ACE-R. The imputed values at death where used in a one-way ANOVA to compare PSPRS and ACE-R between pathology stages. Main effects were considered significant at p < 0.05, we did not correct for multiple comparisons. All statistical analysis and graphical plots were carried out in RStudio (v. 4.0.2).

**Fig. 2.**
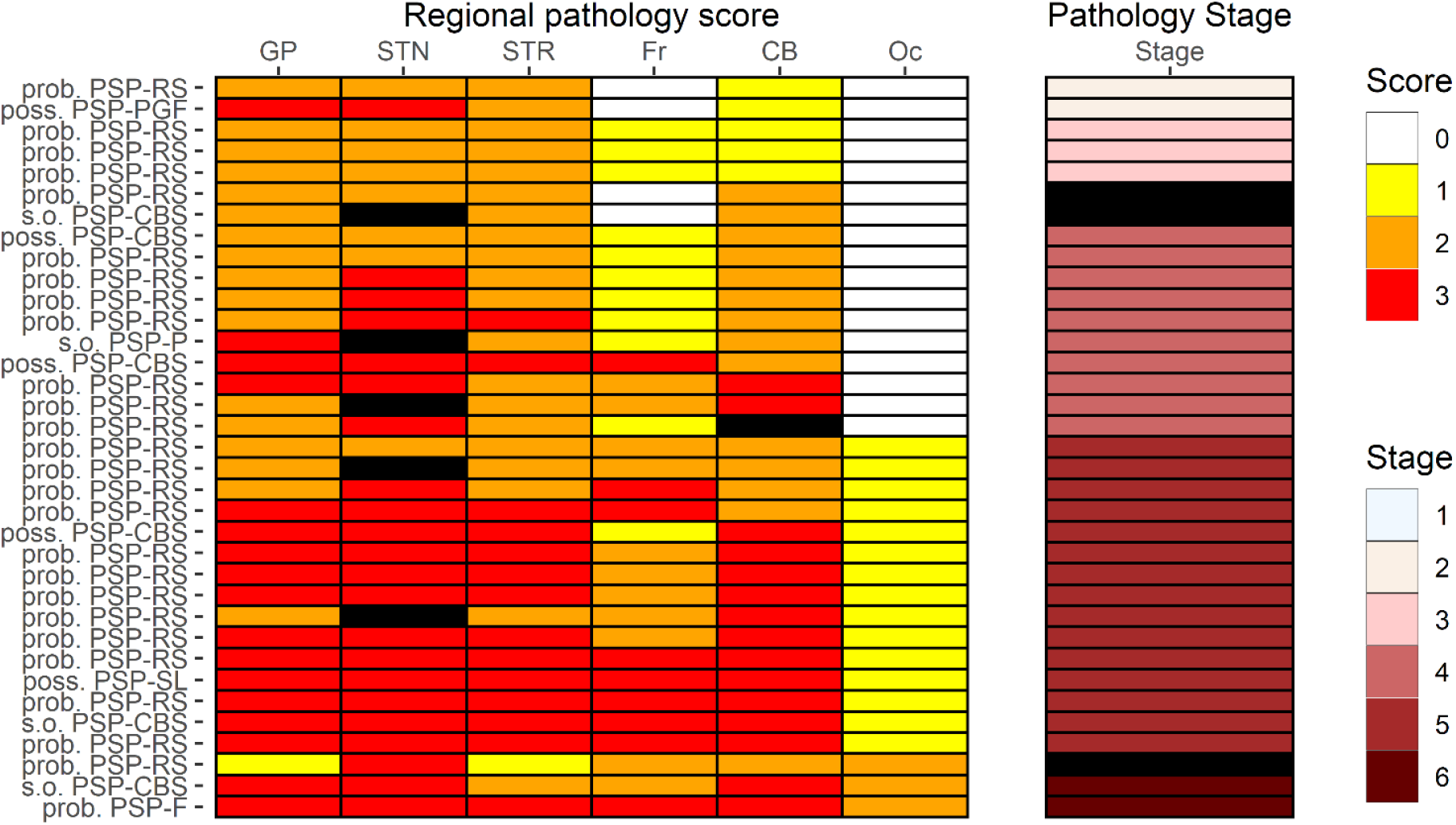
Distribution of pathology scores and pathology stages across 35 PSP cases. Pathology scores are displayed on a colour scale from white – none/0, yellow – mild/1, orange – moderate/2, and red – severe/3. Pathology stages are based on the combination of pathology scores, and are given on a scale from 1-6 (light blue – beige – light pink – coral – dark red – brown). Where a section or region was not available for evaluation, or where the combination of regional pathology scores were incompatible with any of the six pathology stages, the tile is black. For each case the last diagnosis based on the movement disorder criteria is shown to the left of the tiles; PSP – progressive supranuclear palsy, Prob.- probable, poss. – possible, s.o. –suggestive of,PSP-RS-Richardson’s syndrome, PSP-CBS – corticobasal syndrome,PSP-SL – speech/language variant, PSP-F – frontal, PSP-PGF – progressive gait freezing, PSP-P - parkinsonism

## Results

Clinical and demographic details of the 35 cases are summarised in Table 1 and Figure 1. The main group of PSP-RS cases were typical, in their gender balance, mean of age at death of 74, and mean disease duration of 7 years. Variant sub-groups were too small for meaningful statistical comparisons, but note that as expected the male subject with probable PSP-PGF had normal cognition and the male subject with PSP-F had particularly severe cognitive impairments.

We selected 10 random cases and in each area, two authors independently rated tau pathology following Kovacs et al (2020) description, and noting that operational criteria for mild moderate or severe pathology were not reported in their description. In the initial rating, the raters were in agreement in 45/60 regions (75%). Pallidum, cerebellum and occipital lobe had high agreement (≥ 8/10), STN and frontal cortex intermediate (7/10), while STR had low agreement (4/10). This discrepancy was attributed to the lack of operational criteria and different interpretations of the Kovacs et al (2020) ratings for each area, confounded by marked differences between regions in the absolute numbers of immunoreactive cells per field.

We therefore formulated operational criteria with region-specific thresholds for mild pathology as follows: (i) GP and STN mild pathology was defined as 2-10 immunoreactive neurons or oligodendroglia; (ii) in STR, the transition from mild to moderate pathology was marked by the presence of neuronal tau; (iii) frontal and occipital cortex mild pathology was defined as 2-50 tau positive astrocytes; and (iv) cerebellar white matter and/or dentate nucleus mild pathology was defined as 2-50 tau positive oligodendrocytes, neurons or neurofibrillary tangles (Figure 1). A rating of severe pathology was given when one or more tau-positive cells were observed for each each field of view under a 40X objective.

With the new operational criteria the inter-rater agreement increased to 88% (52/59 regions), with high agreement for GP, STR, frontal cortex, occipital and cerebellum (9/10) and intermediate for STN (7/9). Cases were selected at random, with one missing data for STN (hence 59 not 60 regions).

Using these operational criteria 91% (32/35) of cases fell readily into one of the six stages (Figure 2). Of the three cases that did not comply with the staging system, two were clinically probable PSP-RS; one had moderate pathology in cerebellum dentate nucleus and cerebellar white matter but no pathology in frontal cortex, the other had moderate pathology in cortical and cerebellar regions, severe pathology in the subthalamic nucleus, but only mild pathology in the striatum and globus pallidus. A third case, clinically suggestive of PSP-CBS had moderate cerebellar pathology (cerebellum dentate nucleus and cerebellar white matter) but no cortical pathology. The regional distributions of tau pathology across the different clinical subtypes are shown in Figure 3.

**Fig. 3.**
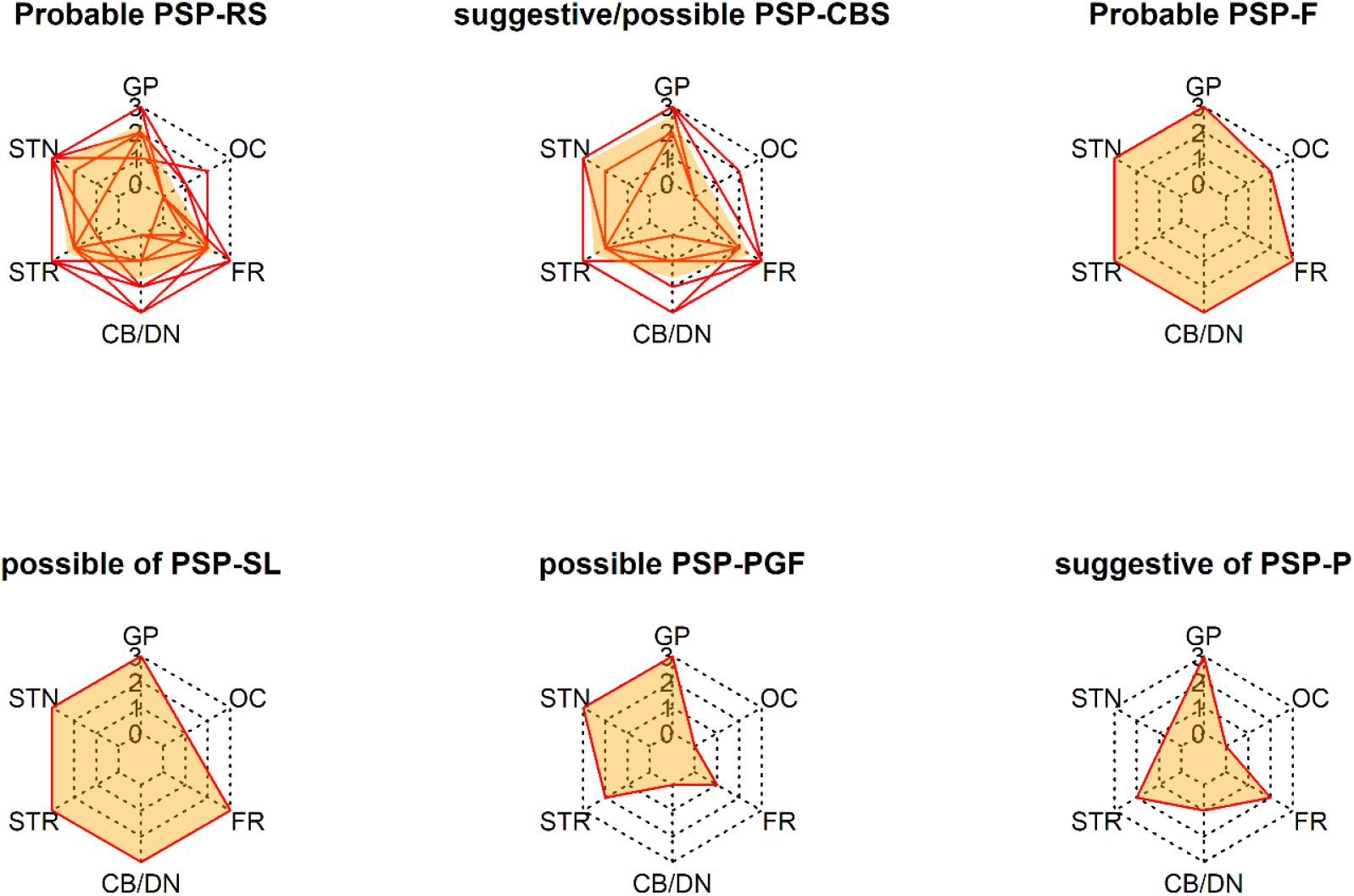
Regional distribution of tau pathology across clinical PSP subtypes. Red lines show individual profiles of regional pathology scores in the globus pallidus (GP), occipital cortex (OC), frontal cortex (FR), cerebellum (CB/DN), striatum (STR), and subthalamic nucleus (STN). Average profiles for probable PSP-RS (n=25) and suggestive off and possible PSP-CBS (n=6) are shown in orange. For possible PSP-PGF, PSP-P, PSP-SL, and PSP-F the single patient profiles are shaded orange. Striped lines show concentric hexagons representing no (0), mild (1), moderate (2) and severe (3) pathology from the innermost to outermost hexagon. PSP – progressive supranuclear palsy, RS – Richardson’s syndrome, CBS – corticobasal syndrome, F-frontal, SL – speech/language, PGF – progressive gait freezing, P – parkinsonism

We then tested whether the pathology staging was associated with demographic, age, and clinical severity. There was no significant association between the pathological stages and age (Kruskal Wallis; n = 32, df = 4, Chi^2^ = 0.80, p = 0.94) or disease duration (Kruskal Wallis; n = 32, df = 4, Chi^2^ = 1.42, p = 0.84).

The interval between death and last assessment of disease severity (PSPRS) and cognitive performance (ACE-R) varied from 24 days to 35 months. To account for the differences in interval between testing and death we took two approaches: (i) correcting for time between last assessment and death (Score ∼ PSP pathology stage | weight = 1 / interval between assessment and death) *and*, (ii) imputing the PSPRS and ACE-R scores at death (imputed score ∼ PSP pathology stage). There were a significant correlation between pathology stage and PSPRS, both when weighting for time between testing and death (F(4,25)=30.0, p = 0.036) and using imputed PSPRS scores (F(4,27)=2.8, p = 0.045)(Fig. 4A and C).

**Fig. 4.**
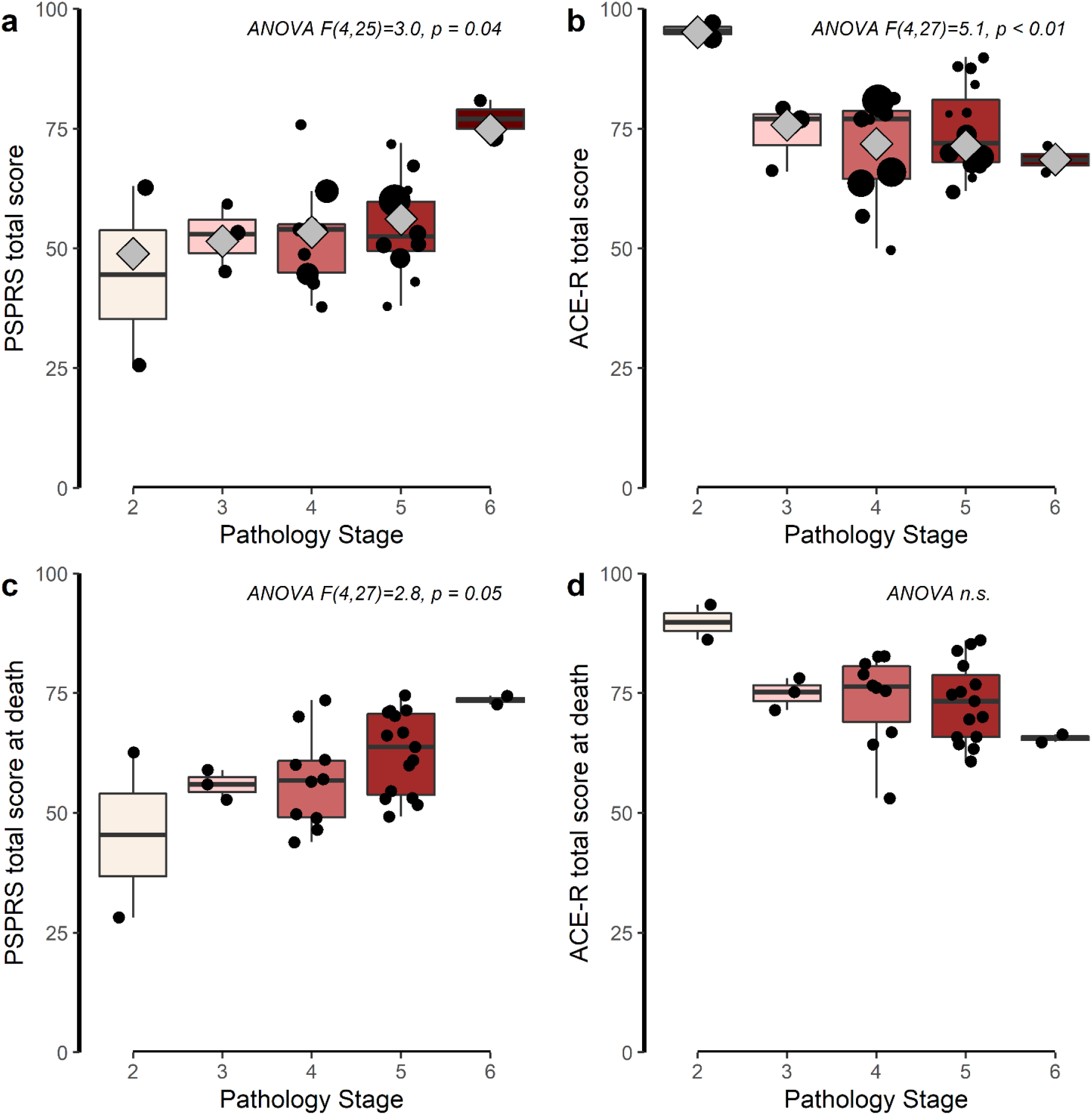
PSPRS and ACE-R according to pathology stage. Individual data points are shown with black dots, boxplots show median with first and third quartiles, extending whiskers to 1.5*interquartile range. A and B, there were a significant effect of pathology stage on PSPRS and ACE-R corrected for the interval between ante mortem assessment and death. Grey diamonds show the weighted means for each group. The size of the point is proportional to the data point’s weight in the analysis. D and E, we imputed the values for PSPRS and ACE-R at death for each individual, black points. There was a significant effect of pathology stage and the imputed values for PSPRS, but not for ACE-R. PSPRS – Progressive Supranuclear Palsy Rating Scale, ACE-R – revised Addenbrooke’s Cognitive Examination, n.s. – not significant, p > 0.05

There was a significant correlation between pathology stage and cognitive deficit, when weighting the analysis for time between testing and death (F(4,27)=5.09, p = 0.0035), but not using imputed ACE-R scores (F(4, 27) = 2.43, p = 0.07) (Fig. 4B and D). Given the small group sizes for stages 2, 3 and 6, *post hoc* analysis were not performed.

## Discussion

The present study provides the first independent validation of the new PSP pathology staging scheme proposed by Kovacs et al. (2020) in an independent cohort. Over 90 % of cases were classifiable into pathology stages 2 to 6. Only three of 35 brains had a combination of regional scores that did not conform to the proposed staging scheme. The disease severity increased with PSP pathology stage, whether using the last PSPRS value before death or imputing a PSPRS value at the time of death based on longitudinal data.

We applied the PSP pathology staging schema, rating tau pathology from none to severe focusing on different cells types in the six regions as described in Kovacs et al. We then defined quantitative operational criteria to distinguish mild from moderate pathology in each region, which was otherwise particularly challenging in the striatum and a major source of disagreement between raters. To improve consistency in striatal ratings we defined a lower absolute threshold for the transition from mild to moderate pathology in the striatum, as the addition of neuronal tau pathology to astrocytic tau. This classification builds on the description of step 2 “A few positive neurons may be seen in the striatum but this is less than the astroglial tau pathology …”[14] but deviates from the simplified approach suggested for the practicing neuropathologist rating only astrocytic pathology in the striatum.

In the regions where neuronal and oligodendroglia tau pathology were rated, moderate pathology in globus pallidus and subthalamic nucleus was defined as more than 10 coiled bodies, neuronal tangles or threads. In our brain series, sections for cortical grey matter areas and cerebellar in which pathology was rated were significantly larger than those for globus pallidus and subthalamic nucleus, and the criteria for moderate was therefore increased to more than 50 astrocytes positive for tau pathology. Applying our region-specific thresholds for rating of tau pathology, 32 out of 35 brains conformed to the proposed staging scheme, including ten with diverse clinical phenotypes. All of our cases had clinical diagnosis of PSP (probable, possible or suggestive of) which may explain why we lacked cases in stage 1, and had only a few in stage 2 and 3. This lack of a floor effect in staging clinically diagnosed cases supports the potential utility of the staging system to identify early and pre-symptomatic cases of PSP.

Defining the criteria for pathological ratings is a challenge faced by all pathological staging systems, aiming for both ease of use and consistency across time and raters [29]. This is especially true for rare disease where a neuropathologist may not have seen many cases. For staging of PSP pathology Williams et al. [30] used the distribution of total counts of tau pathology (coiled bodies and threads) to determine region specific thresholds for grades 0-4 (none to severe) and provided a visual guide for rating of pathology. They report a high agreement between two independent neuropathologists (weighted kappa 0.71), but this staging system is not in common clinical use. In their analysis the tau score of substantia nigra, caudate and dentate nucleus was the preferred surrogate marker for total tau pathology, with minimal pathology being defined as tau aggregations in subthalamic nucleus, substantia nigra and globus pallidus. Their staging scheme shows a differential progression of tau pathology in caudate and putamen, where the caudate progresses from none to grade 2, and putamen remains at grade 1. In our study the putamen, globus pallidus and subthalamic nucleus were evaluated in one section taken just the posterior portion to the mammillary bodies. At this level the caudate is small and located too dorsally to be included. Thus, we have most likely assessed pathology in the putamen posterior to the level of Williams et al. and Kovacs et al. who assessed the caudate and putamen on the same section, and subthalamic nucleus in a separate section.

We speculate that differences in sampling site along the rostro-caudal axis may have contributed to our lower average rating of striatal pathology compared to Kovacs et al. [14]. We do not question the significance of striatal pathology in PSP, but highlight the challenges arising from the inclusion of striatum in the PSP staging scheme: *first*, total tau pathology scores in striatum does not differ between clinical subtypes [14]; *second*, astrocytic tau pathology only differs significantly between PSP-P and PSP-CBS [14]; and *third*, the single scenario where rating of tau pathology in striatal astrocytes would distinguish between two stages, 1 and 2, is if pathology in the globus pallidus and the subthalamic nucleus is rated moderate, no pathology is observed in frontal, cerebellar and occipital regions, and striatal pathology is mild – this combination was not observed in the 206 cases assessed by Kovacs et al. nor in the 35 cases assessed here.

Half of our cases had higher pathology ratings in dentate/cerebellum than frontal cortex, making them “cerebellar dominant” while only 8% of cases were frontal predominant. This contrasts with Kovacs et al. who observed nearly equal numbers of cerebellar and frontal dominant cases in their PSP-RS group. However, in the 34 cases review by Williams et al. [30], ratings of tau pathology in cerebellum were on average higher than frontal cortex. The small number of patients in PSP-CBS and single patients other clinical subtypes prevented us from testing the regional distribution of tau pathology between groups. However, our observations are in agreement with previous studies with all subtypes having moderate to severe pathology in basal ganglia structures and variable pathology in cortical and cerebellar region: with high (moderate to severe) cerebellar and frontal tau pathology in PSP-CBS, PSP-SL and PSP-PF[14, 16, 25], and milder (none to moderate) tau pathology in cortical and cerebellar regions in PSP-P and PSP-PGF [1, 14, 23, 24, 30]. Differences in proportion of frontal versus cerebellar predominance may therefore, in part, be explained between the proportion of different clinical subtypes between the present study, Kovacs et al. [14] and Williams et al. [30].

In addition to the validation of the new pathology staging system, and the suggested new operational criteria, a key result from our study is the significant association between pathology stage and clinical severity - measured by PSPRS or cognition. The PSPRS is a composite score (0-100 points) of the severity of symptoms and signs across six domains: behaviour, gait, mobility, bulbar, limb, and oculomotor problems [9]. PSPRS scores were higher in cases with higher PSP pathology stages, both for imputed and nearest-death score. Correspondingly, a recent study showed correlations between the presence of behavioural, executive, motor and visuospatial deficits and *post mortem* FTD-tau scores collated from glial and neuron pathology in 19 regions [24], supporting the link between the extend of tau pathology spread and clinical severity in PSP. We report a significant correlation between ACE-R scores nearest to death and PSP pathology stage. This supports a previous association between cognitive impairment and pathological tau in PSP [13].

A limitation of our study is the modest sample size, particularly of phenotypes other than PSP-RS. A significant strength of our study is the prospective collection of longitudinal clinical data. However, due to the nature of *post mortem* studies, time intervals between last *ante mortem* data collection and death varied widely between cases. To overcome this limitation we adjusted for the time interval in our analysis, taking advantage of the longitudinal data available for most patients in order to impute the scores at death [18].

Overall our study supports the validity of the proposed PSP pathology staging system, being easy to implement in the day-to-day neuropathological evaluation (and retrospectively) as the regions required are routinely sampled for pathological diagnoses of neurodegenerative diseases. We show that the proposed PSP staging schema is applicable across the spectrum of clinical PSP subtypes with >90% of cases fulfilling staging criteria. In addition to the written description provided by Kovacs et al. we suggest region-specific quantitative criteria along with a visual guide for the rating of tau pathology in the six regions. Together with high compliance with the staging scheme, our findings suggests that the sequential distribution of tau pathology is associated with progressive clinical severity in PSP.

## Data Availability

The datasets used are available from the corresponding author on reasonable request.

## Acknowledgements

The authors would like to thank the research participants and caregivers, and the staff at the Cambridge Centre for Parkinson-Plus and the Department of Histopathology and Cytology at the Cambridge Brain Bank.

## Availability of data and materials

The datasets used are available from the corresponding author on reasonable request.

## Authors’ contribution

All authors contributed to the study conception and design. Material preparation, data collection and analysis were performed by Mayen Briggs, Kieren Allinson, Maura Malpetti, and Sanne Simone Kaalund. James Rowe and Maria Spillantini critically revised and contributed important intellectual content to the manuscript. All authors read and approved the final manuscript.

## Consent for publication

Not applicable.

## Ethics approval and consent to participate

The study ethics was approved by the Health Research Authority, NHS, England (IRAS-202 802, “Neurodegeneration Research in Dementia”). The PiPPIN (Pick’s Disease and Progressive Supranuclear Palsy: Prevalence and Incidence) Study was approved by Cambridge’s research ethics committee. The study was conducted in accordance with the 1964 Helsinki declaration.

