## Supplementary Figures 1 and 2 for "Validation of the new pathology staging system for progressive supranuclear palsy"

### Title

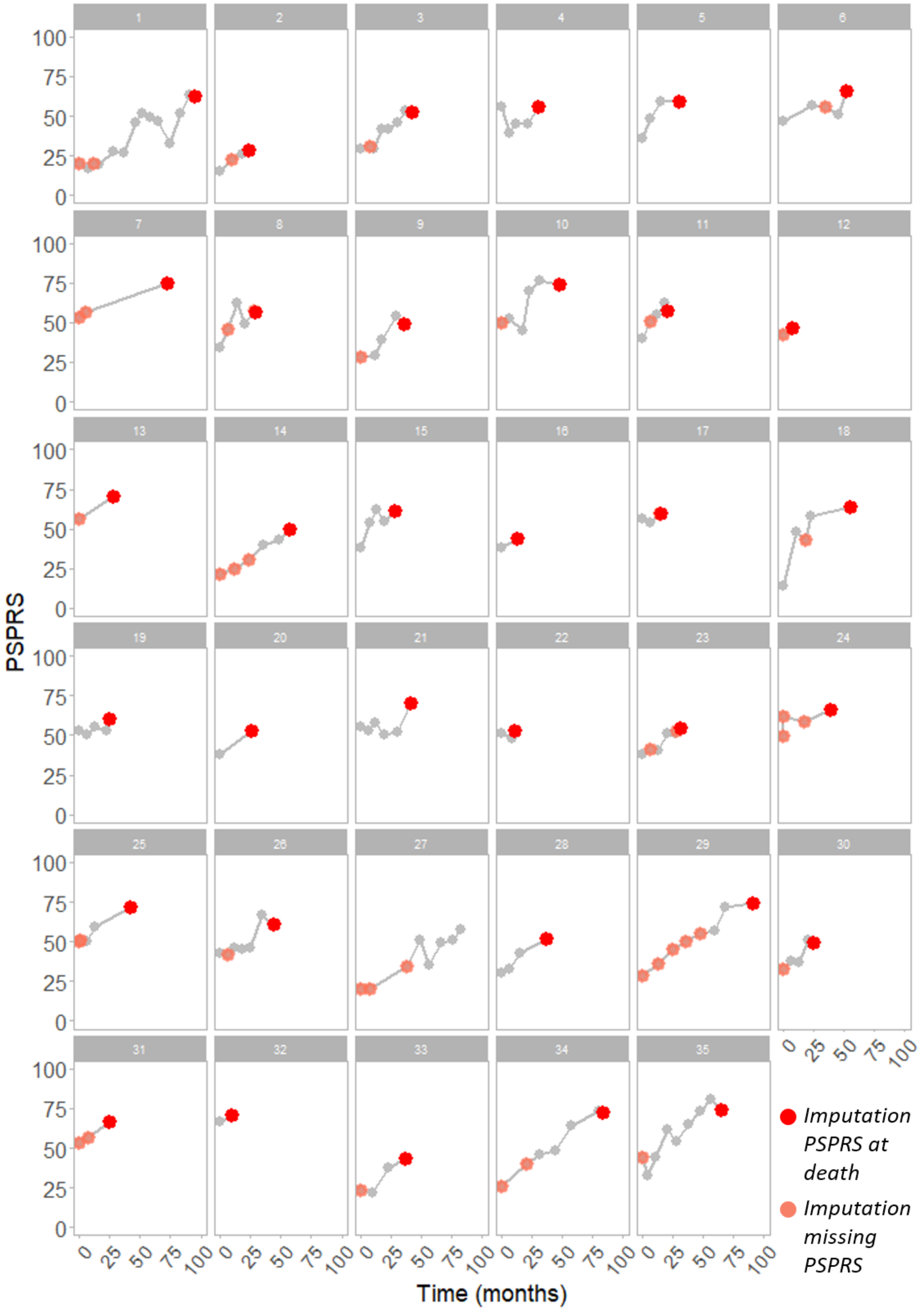


***Supplementary Fig. 1*** *Longitudinal and imputed PSPRS for individual PSP patients. PSPRS and ACE-R accessed at visits to the clinic (grey points) were used to impute PSPRS values at death (red points) and at visits where only ACE-R were measured (pink points) for each of the 35 PSP patients. The x-axis show time from the first visit (0) to death in months. The order of patients (1-35) is commensurate with that of Figure 2. ACE-R – revised Addenbrooke's Cognitive Examination revised, PSPRS – Progressive Supranuclear Palsy Rating Scale*

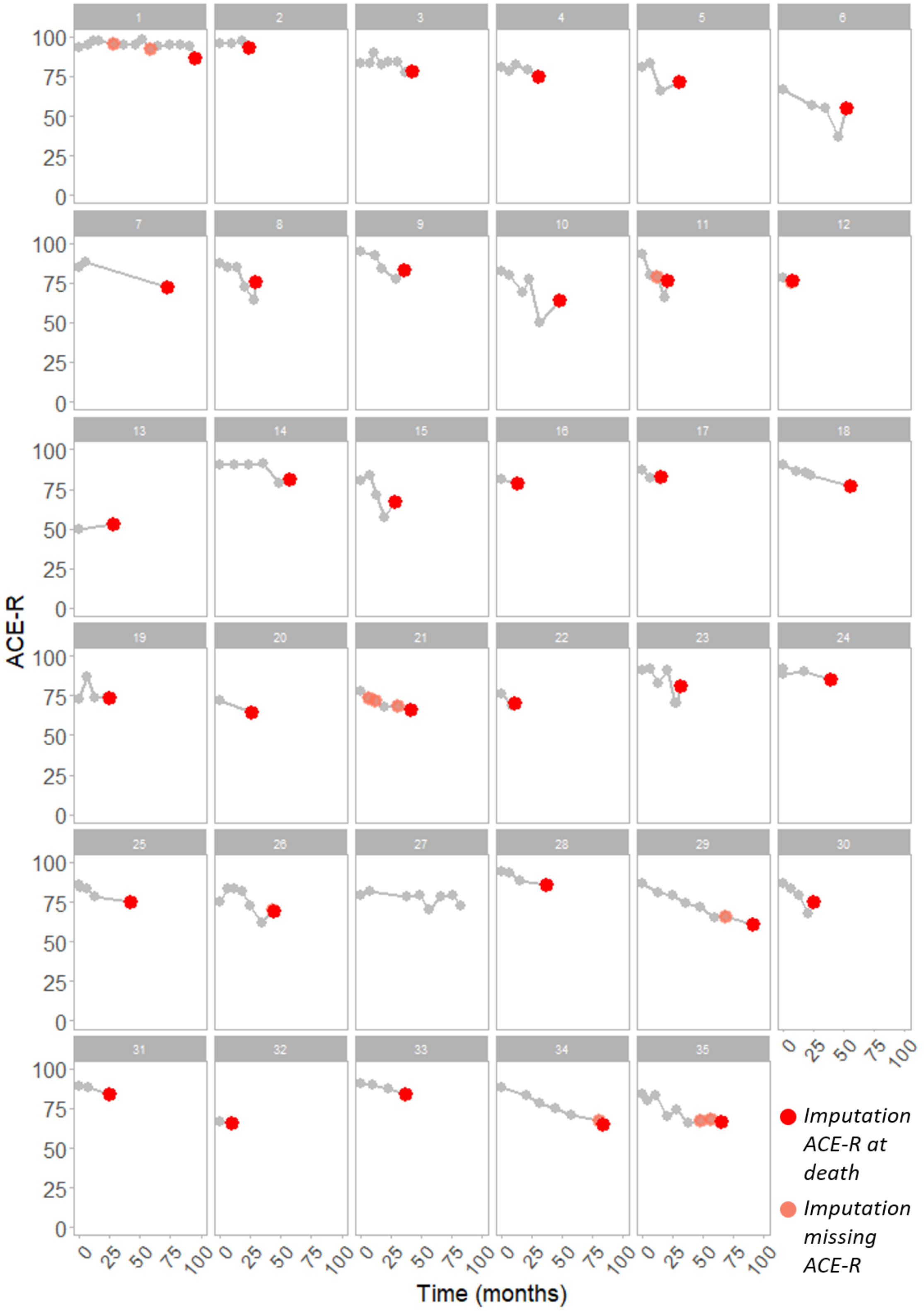


***Supplementary Fig. 2*** *Longitudinal and imputed ACE-R for individual PSP patients. PSPRS and ACE-R accessed at visits to the clinic (grey points) were used to impute ACE-R values at death (red points) and at visits where only PSPRS were measured (pink points) for each of the 35 PSP patients. The x-axis show time from the first visit (0) to death in months. The order of patients (1-35) is commensurate with that of Figure 2. ACE-R – revised Addenbrooke's Cognitive Examination revised,* *PSPRS – Progressive Supranuclear Palsy Rating Scale*
